# Utility of estimated pulse wave velocity for assessing vascular stiffness: comparison of methods

**DOI:** 10.1101/2022.01.13.22268898

**Authors:** Stefan Möstl, Fabian Hoffmann, Jan-Niklas Hönemann, Jose Ramon Alvero-Cruz, Jörn Rittweger, Jens Tank, Jens Jordan

## Abstract

**Aim:** Pulse wave velocity independently predicts cardiovascular risk. Easy to use single cuff oscillometric methods are utilized in clinical practice to estimate pulse wave velocity. We applied the approach in master athletes to assess possible beneficial effects of lifelong exercise on vascular health. Furthermore, we compared single cuff measurements with a two-cuff method in another cohort.

**Methods:** We obtained single cuff upper arm oscillometric measurements thrice in 129 master athletes aged 35 to 86 years and estimated pulse wave velocity using the ArcSolver algorithm. We applied the same method in 24 healthy persons aged 24 to 55 years participating in a head down tilt bedrest study. In the latter group, we also obtained direct pulse wave velocity measurements using a thigh cuff.

**Results:** Estimated pulse velocity very highly correlated with age (R^²^= 0.90) in master athletes. Estimated pulse wave velocity values were located on the same regression line like values obtained in participants of the head down tilt bed rest study. The modest correlation between estimated and measured PWV (r^2^ 0.40; p<0.05) was attenuated after adjusting for age; the mean difference between pulse wave velocity measurements was 1 m/s.

**Conclusion:** Estimated pulse wave velocity mainly reflects the entered age rather than true vascular properties and, therefore, failed detecting beneficial effects of life long exercise.

**Funding:** The AGBRESA-Study was funded by the German Aerospace Center (DLR), the European Space Agency (ESA, contract number 4000113871/15/NL/PG) and the National Aeronautics and Space Administration (NASA, contract number 80JSC018P0078). FH received funding by the DLR and the German Federal Ministry of Economy and Technology, BMWi (50WB1816). SM, JT and JJ were supported by the Austrian Federal Ministry for Climate Action, Environment, Energy, Mobility, Innovation and Technology, BMK (SPACE4ALL Project, FFG No. 866761).

## Main Text

Aortic pulse wave velocity (PWV) which relates to vascular stiffness, independently predicts cardiovascular risk including stroke ^1, 2^. PWV values above 10 m/s have been suggested as threshold indicating increased risk ^3^ and are included in current hypertension guidelines for assessing hypertension-mediated organ damage and guiding secondary prophylaxis ^4^. However, these guidelines do not specify certain methods to measure PWV. Because measurements at different sites including the thigh are required, PWV-assessment is difficult to implement in busy clinics or in large scale studies. Oscillometric blood pressure monitors estimating PWV based on age and blood pressure could simplify vascular assessment ^1, 5^. Indeed, estimated PWV correlates with age ^6^ and invasively measured PWV ^1^. These monitors operate observer-independent, are easy to use, and are, therefore, applied in clinical practice and in clinical studies alike ^7^. However, in a study investigating beneficial effects of competitive life-long physical exercise in elite Masters Athletes, we obtained estimated PWV results suggesting that the approach adds little information to classical cardiovascular risk assessment. We decided comparing the methodology with aortic PWV measurements in an independent set of healthy persons participating in a head-down-tilt bedrest study.

All subjects provided informed consent before enrollment. Studies were approved by the Northrine-Medical-Association (Ärztekammer Nordrhein) ethics committee and registered at the German Clinical Trial Register (DRKS00015677 and DRKS00015172). According to Kolmogorov-Smirnov-Test, all data were normally distributed and results are reported as mean values ± standard deviation. Based on other cardiovascular risk-estimate-models, we used a quadratic regression model.

During the 23^rd^ Masters Athletics Championships 2018 in Málaga, Spain, we approached 163 master athletes. We excluded athletes with atrial fibrillation or significant cardiovascular disease assessed by echocardiography. In the remaining 129 athletes (88 men/41 women, 56±11 (range 35-86) years, 24.0±3.5 kg/m^2^), we acquired blood pressure, heart rate, and estimated PWV thrice on the same arm after 10 minutes supine rest (Arc-Solver-Algorithm, CardioCube, AIT, Vienna, Austria). Resting heart rate was 61±11 bpm and blood pressure was 128±15/78±8 mmHg. Estimated PWV ranged from 5.5-14.5 with a mean of 8.3±1.8 m/s. In a quadratic regression model, age and mean arterial pressure predicted estimated PWV (beta-value 0.93 and 0.14, *p*<0.001), whereas sex and BMI had no influence. The model explained 95% (R^2^=0.95) of estimated PWV’s variance. Age alone explained 90% (R^2^=0.90).

In 24 healthy participants (16 men/8 women, 33±9 years, 24.3±2.1 kg/m^2^) of the AGBRESA (artificial gravity bedrest) study, which was conducted in collaboration with NASA and ESA in Cologne, Germany ^8^, we assessed estimated PWV before head-down-tilt bedrest as described above. As the estimated PWV has been validated against invasive catheter measurements derived from the ascending aorta to the aortic bifurcation ^1^, we mimicked this approach non-invasively by measuring pulse wave arrival time from the electrocardiographic R-Peak to the arrival of the pulse wave at an oscillometric thigh-cuff. We corrected pulse wave arrival time for isovolumetric contraction time assessed by 2D-Pulsed-Wave-Doppler echocardiography. Dividing jugulum-thigh cuff distance by corrected pulse wave arrival time resulted in measured PWV.

Resting heart rate was 62±9 bpm and blood pressure was 125±11/70±7 mmHg. Estimated PWV was 5.8±1.1 m/s and measured PWV was 4.8±0.6. Age and estimated PWV were highly correlated (R^2^ 0.88, p<0.001, Figure 1A). The correlation between measured PWV and age was weaker (R^2^ 0.55, p<0.001). In a quadratic regression model, age (beta-value 0.99, *p*<0.001) but not body-mass-index, sex, or mean arterial pressure predicted estimated PWV. The model explained 98% of estimated PWV’s variance. R^2^ between estimated and measured PWV was 0.40 (p<0.05). We observed an increasing bias between estimated and measured PWV with advancing age with an average 0.96±0.83 m/s difference between methods (Figure 1B). When adjusting the correlation of measured and estimated PWV for age we could no longer observe a significant relationship (*p*=0.267).

**Figure 1 A-C.**
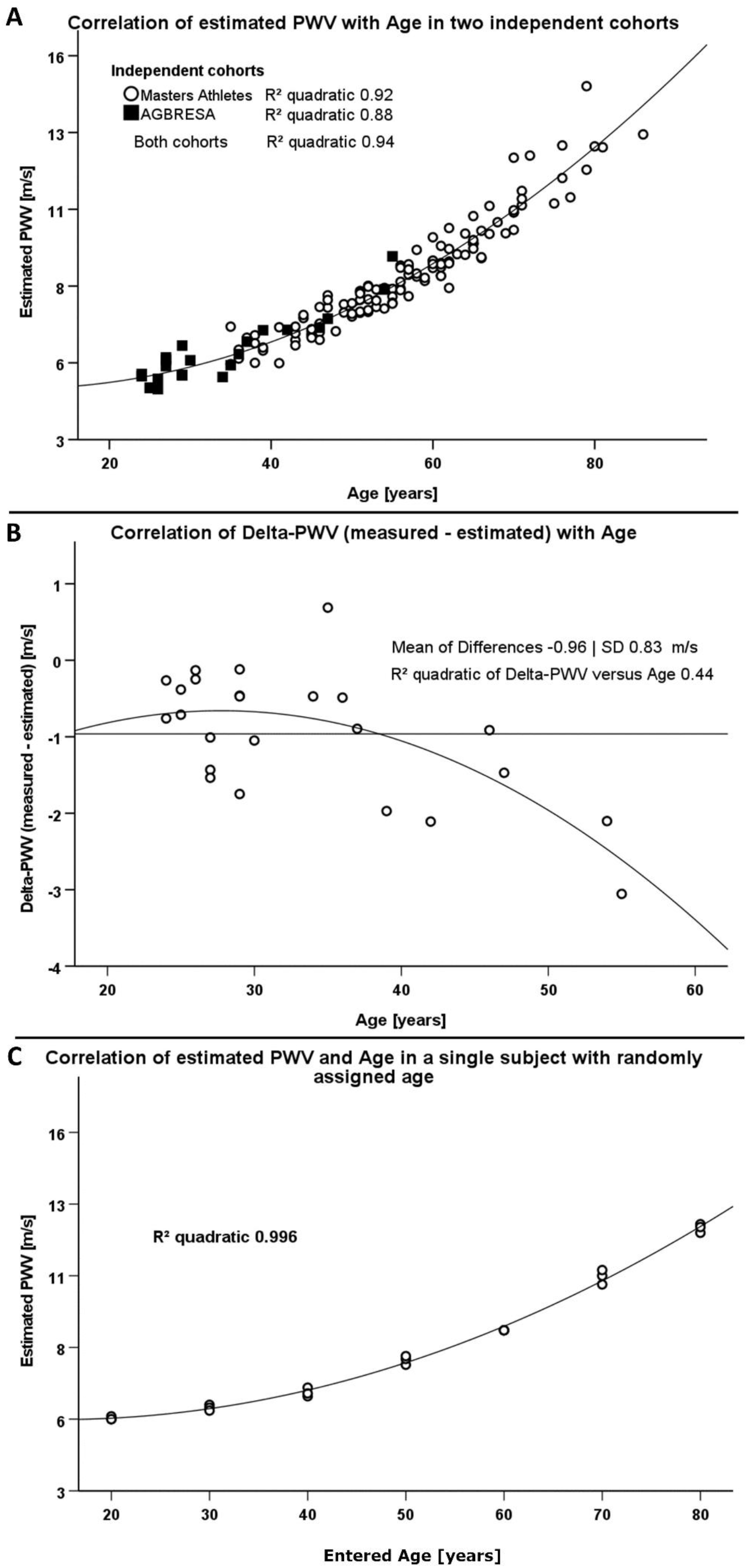
Comparison of two different methodologies to assess pulse wave velocity (PWV) in two different cohorts. 1A: Correlation between estimated PWV and age in 129 Masters Athletes (empty circles) and in 24 AGBRESA-study participants (black squares). 1B: Regression analysis between the measured PWV and estimated PWV difference with age in 24 AGBRESA-study participants. Estimated deviated from measured PWV with increasing age (R^2^ 0.44, p=0.07). 1C: Quadratic regression analysis between estimated PWV and entered age in one man being in his 50s. We randomly entered age in decades from 20-80 and obtained measurements thrice at each entered age.

To assess influences of entered rather than chronological age on estimated PWV, we repeated measurements in one male person, who is in his 50s and has normal BMI, and randomly changed the entered age in decades from 30-80 years. We obtained measurements in each entered age decade thrice. Again, estimated PWV related to entered age (R^2^ 0.996, *p*<0.001, figure 1C) akin to the relationship between estimated PWV and chronological age in athletes and in bedrest-study participants.

The idea of obtaining vascular measurements, such as PWV, is gaining insight in individual vascular risk above and beyond traditional risk factors. The clinical goal is to target preventive measures to patients most likely to benefit. Similarly to other studies^6^, we observed an almost perfect correlation between estimated PWV and age in two independent cohorts. Strikingly, estimated PWV in elite master athletes, which would be expected to benefit from life-long exercise, and in healthy bedrest study participants of average physical fitness were located on the same regression line. Suffice it to say that many athletes were frustrated when we reported their findings.

Theoretically, our findings could result from methodological limitations of PWV estimates or a rather limited effect of lifelong exercise on aortic stiffness. A previous study showed significantly lower measured PWV in endurance trained compared to sedentary older men ^9^ making a methodological limitation more likely. Indeed, in our study, estimated PWV substantially overestimated PWV, particularly in older persons. Moreover, the modest correlation between estimated and measured PWV was attenuated after adjusting for age in our relatively small bedrest cohort. Finally, in a previous study, estimated PWV highly correlated with PWV calculated solely from age and blood pressure ^5^. Thus, the algorithm providing PWV estimates appears to weigh age so strongly that subtle influences on vascular wall properties caused by hemodynamic changes and physical training cannot be discerned. Even more so, estimated PWV mainly reflects data entered prior to measuring blood pressure rather than true vascular properties. The clinical implication is that estimated PWV is no substitute for measured PWV, which likely limits the utility the methodology in individualizing risk assessment. In fact, simply asking the patient in front of us for his or her age and measuring blood pressure provides almost as much information as estimating PWV. Instead, vascular ageing assessment using established methodologies rather than estimates should be considered ^10^.

## Data Availability

The raw data supporting the conclusions of this article will be made available by the authors upon request, without undue reservation.

## Acknowledgements

We would like to thank Siegfried Wassertheurer, Bernhard Hametner and Martin Bachler from the Austrian Institute of Technology for providing the CardioCube.

## Disclosures

We declare no conflict of interest.

## Notes

### Competing Interest Statement

The authors have declared no competing interest.

### Clinical Trial

DRKS00015677 and DRKS00015172

### Funding Statement

The entire AGBRESA-Study was funded by the German Aerospace Center (DLR), the European Space Agency (ESA, contract number 4000113871/15/NL/PG) and the National Aeronautics and Space Administration (NASA, contract number 80JSC018P0078). Fabian Hoffmann received funding by the German Aerospace Center (DLR) and the German Federal Ministry of Economy and Technology, BMWi (50WB1816).

### Author Declarations

Studies were approved by the Northrine-Medical-Association (Aerztekammer Nordrhein) ethics committee.

## References

1. Hametner B, Wassertheurer S, Kropf J, Mayer C, Eber B, Weber T. Oscillometric estimation of aortic pulse wave velocity: Comparison with intra-aortic catheter measurements. Blood pressure monitoring. 2013;18:173–176

2. Vishram-Nielsen JKK, Laurent S, Nilsson PM, Linneberg A, Sehested TSG, Greve SV, Pareek M, Palmieri L, Giampaoli S, Donfrancesco C, Kee F, Mancia G, Cesana G, Veronesi G, Kuulasmaa K, Salomaa V, Kontto J, Palosaari T, Sans S, Ferrieres J, Dallongeville J, Söderberg S, Moitry M, Drygas W, Tamosiunas A, Peters A, Brenner H, Njolstad I, Olsen MH. Does estimated pulse wave velocity add prognostic information?: Morgam prospective cohort project. Hypertension. 2020;75:1420–1428

3. Van Bortel LM, Laurent S, Boutouyrie P, Chowienczyk P, Cruickshank JK, De Backer T, Filipovsky J, Huybrechts S, Mattace-Raso FU, Protogerou AD, Schillaci G, Segers P, Vermeersch S, Weber T. Expert consensus document on the measurement of aortic stiffness in daily practice using carotid-femoral pulse wave velocity. J Hypertens. 2012;30:445–448

4. Williams B, Mancia G, Spiering W, Agabiti Rosei E, Azizi M, Burnier M, Clement DL, Coca A, de Simone G, Dominiczak A, Kahan T, Mahfoud F, Redon J, Ruilope L, Zanchetti A, Kerins M, Kjeldsen SE, Kreutz R, Laurent S, Lip GYH, McManus R, Narkiewicz K, Ruschitzka F, Schmieder RE, Shlyakhto E, Tsioufis C, Aboyans V, Desormais I. 2018 esc/esh guidelines for the management of arterial hypertension: The task force for the management of arterial hypertension of the european society of cardiology and the european society of hypertension. J Hypertens. 2018;36:1953–2041

5. Hametner B, Wassertheurer S, Mayer CC, Danninger K, Binder RK, Weber T. Aortic pulse wave velocity predicts cardiovascular events and mortality in patients undergoing coronary angiography: A comparison of invasive measurements and noninvasive estimates. Hypertension. 2021;77:571–581

6. Schwartz JE, Feig PU, Izzo JL, Jr. Pulse wave velocities derived from cuff ambulatory pulse wave analysis. Hypertension. 2019;74:111–116

7. Paiva AMG, Mota-Gomes MA, Brandão AA, Silveira FS, Silveira MS, Okawa RTP, Feitosa ADM, Sposito AC, Nadruz W, Jr. Reference values of office central blood pressure, pulse wave velocity, and augmentation index recorded by means of the mobil-o-graph pwa monitor. Hypertension research : official journal of the Japanese Society of Hypertension. 2020;43:1239–1248

8. Hoffmann F, Rabineau J, Mehrkens D, Gerlach DA, Moestl S, Johannes BW, Caiani EG, Migeotte PF, Jordan J, Tank J. Cardiac adaptations to 60 day head-down-tilt bed rest deconditioning. Findings from the agbresa study. ESC heart failure. 2020;8:729–744

9. Vaitkevicius PV, Fleg JL, Engel JH, O’Connor FC, Wright JG, Lakatta LE, Yin FC, Lakatta EG. Effects of age and aerobic capacity on arterial stiffness in healthy adults. Circulation. 1993;88:1456–1462

10. Jordan J, Nilsson PM, Kotsis V, Olsen MH, Grassi G, Yumuk V, Hauner H, Zahorska-Markiewicz B, Toplak H, Engeli S, Finer N. Joint scientific statement of the european association for the study of obesity and the european society of hypertension: Obesity and early vascular ageing. J Hypertens. 2015;33:425–434

